# Determinants of dietary salt intake in adults: A protocol for Scoping Review

**DOI:** 10.1101/2021.08.15.21262091

**Authors:** Raunaq Singh Nagi, Pankaj Prasad, Sanjeev Kumar

## Abstract

**Objective:** The objective of this scoping review is to methodically review the current literature and identify the factors/determinants of dietary salt consumption in adults.

**Introduction:** High dietary salt intake has been identified as a risk factor for non-communicable diseases and conditions. Despite continuous and rigorous efforts, dietary salt intake still remain above the recommended adult daily dose of 5g, both locally and globally. This indicates existence of unidentified or unaddressed behavioural factors that diminish efficiency of salt reduction efforts targeted towards public.

**Inclusion Criteria:** This review will include global literature dealing with factors associated with dietary salt intake in adults. Qualitative, quantitative and ecological studies on behavioural, psychosocial and environmental factors associated with awareness regarding dietary salt intake and barrier to its reduction will be considered. Studies published only in English language, without any limits on date of publication will be considered for this review.

**Methods:** A comprehensive search across databases namely, MEDLINE, ScienceDirect, JSTOR, ERIC, DOAJ and OATD will be carried out to retrieve and identify relevant literature. Two reviewers will screen the titles followed by abstracts and subsequently full-length texts according to the inclusion and exclusion criterion, removing unrelated studies and finally compiling and extracting information from chosen studies in data extraction forms. Descriptive statistics will be used to represent the data. Thematic analysis of extracted data using deductive approach will be conducted.

## Introduction

Excessive dietary intake of salt or sodium has been identified as a behavioural risk factor for incidence and progression of a number of non-communicable diseases (NCDs) ranging from gastric cancer (1) to cardiovascular diseases. (2) Evidence regarding adverse effects of high salt consumption started accumulating in the mid-twentieth century(3), and several guidelines have been formulated by the World Health Organization (WHO) for cautious use and reduction of dietary salt under the daily adult recommended dosage of 5g/day. (4) To reduce the global mortality by 25% WHO unveiled the Global Action Plan for Prevention and Control of NCDs in 2013 enlisting policy level recommendations to initiate responses to achieve a dietary salt intake reduction of 30% at population level by 2025. (5) Several countries, including India have issued national guidelines and formulated policies to complement WHO’s global action. (6) Despite these prompt efforts, reduction in dietary salt intake has not been achieved to desirable levels at population level and salt intake remains high across the globe. (6–9)

This scoping review is being undertaken to identify the factors associated with consumption of high quantities of dietary salt at individual, household and community levels. This study would also attempt to identify the barriers of salt reduction in individuals who are aware of the risks of high salt consumption. For the purpose of generating a logic map of factors of dietary salt intake two theoretical models, namely, Health Belief Model (10) and Transtheoretical Model (11) would be used to understand and chart the behaviours and practices pertaining to salt consumption at all the three levels.

This evidence will aid in formulation a theoretical framework for development of appropriate and targeted behavioural change interventions for the global aim of population level salt reduction. For this review we are using freely searchable databases only, which might be a potential limitation of the study. We are employing lot quality assurance in addition to kappa statistics to obtain maximum quality control in the review. We have not observed this approach being used in any other scoping review yet. Additionally, to the best of our knowledge, this is the first study being undertaken to identify the determinants of dietary salt intake in adults. The protocol will result in synthesis of evidence regarding the perceptions of adult population about the sources, their awareness regarding the effects of excessive dietary salt, social factors, environmental factors, and practices that lead to intake of higher than recommended quantities of dietary salt, and barriers in reducing dietary salt and compliance with salt reduction guidelines.

## Review Questions

The primary research question of the study has been identified as “What are the factors which influence consumption of salt in adults at individual, household, and community level?” And the aim of the study is “To identify factors associated with dietary salt consumption in adults”.

To achieve this, the following questions will be used to determine various aspects of salt consumption by adults:

a. What are the factors which influence the knowledge regarding consumption of salt with human health?
b. Why people are not concerned about the harmful effects of excess salt consumption?
c. What is the awareness, if any, regarding the sources and effects of high salt intake in the population?
d. What are the practices that contribute to high intake of salt?
e. What are the barriers in reducing dietary salt in individuals and households that are aware of the risks associated with salt intake?

## Inclusion Criteria

### Participants

This review will inspect community-based studies on adults aged ≥18 years regardless of their gender, ethnicity or origin. We would exclude any study which focus only on a sub-population of adults, such as any particular disease or disorder.

### Concept

The concepts being dealt with in this study include the factors which determine the knowledge, attitudes, behaviours, beliefs and/or practices of adults that govern dietary salt intake in adults. This will also be inclusive of the barriers which render adults who are aware of high salt incapable of reducing their dietary salt intake.

### Context

This review will consider all the primary data analyses, but not risk factor prevalence surveys or facility-based studies. Studies that deal with one or the other factor which leads to behavioural modulation will be included while guidelines and recommendations will be excluded.

### Study Type

This review will include qualitative, quantitative or mixed methods studies, and cohort and cross-sectional, randomized controlled trials type study designs will be included. Additionally, ecological studies, especially the ones focussing on built environment will be included.

Studies will be excluded if they are editorials, opinions, commentaries, perspectives, guideline and recommendations, viewpoints, book chapters, policy level interventions without consideration of behaviour change. Similarly, case reports and case series will be excluded for low confidence of evidence. Also, studies in which ALL the authors are on payroll of pharmaceuticals/nutraceuticals will not be included.

## Methods

This review follows the methodology of scoping study as outlined by Arksey and O’Malley (12) and scoping framework recommendations of Joanna Briggs Institution. (13)

Preferred Reporting Items for Systematic reviews and Meta-Analyses extension for Scoping Reviews (PRISMA-ScR) guidelines and checklist will be used for reporting the findings of the study. (14)

### Search Strategy

The initial search has been conducted using keywords like, “adult”, “determinant”, “dietary salt intake”, “dietary sodium intake” and subsequently other relevant keywords were identified by tracking the titles, abstracts and subject headings (such as MeSH in Medline) and included in the search. Appropriate search builder tools (Boolean operators for MEDLINE, JSTOR, DOAJ and ScienceDirect or, a combination of Boolean operators and mathematical symbols such as “+” and “-” for ERIC) were used and various filters and limits, such as for the type of study and language were applied. Permutated combinations and variations of these keywords were used to arrive at the final search term and strategy, customized for each of the databases. The search terms and strategy were tested, iterated and optimized for individual databases. After the preliminary search, a penultimate search was conducted from 23 July 2020 to 30 July 2020 across all the databases. The final search strategy employed for the conducting this search on three of the databases has been provided in Appendix I. A final search will be carried out and search results of the databases will be either exported or copied in “.txt” (for ScienceDirect, JSTOR and ERIC) or “.csv” (for Medline, DOAJ and OATD) formats as per the availability on the database. Final results obtained will be tabulated in Microsoft Excel 2019 (Redmond, Washington, USA) saved in “.xlsx” format. After identification of relevant studies, citation tracking and snowballing will be used to identify more studies to be included for later stage of analysis.

### Information sources

For the purpose of identification of studies, only the freely searchable databases are chosen. The following databases are included in the study: MEDLINE, ScienceDirect, JSTOR, ERIC, DOAJ. For retrieval of masters and doctoral theses, Open Access Thesis Database (OATD) has been chosen.

### Study Selection

Two reviewers will independently screen the titles and subsequently the abstracts of the studies in accordance to the inclusion and exclusion criteria. In stage I, each and every title of the studies retrieved from all the databases will be screened and excluded or included as per the eligibility criteria. Abstracts of all the studies included for stage II evaluation will be retrieved, screened and sorted as per the eligibility criteria. Studies for which abstracts are unavailable will be excluded. After omission of studies at stage II, full length articles will be retrieved and assessed for relevance. Studies for which full-length articles are unavailable freely will be excluded. After the selection of articles for full length screening, stage III, any ambiguity in the eligibility of full text articles will be resolved by the help of a third independent reviewer. Any other studies identified to be of relevance during full length screening of the previously screened articles will also be included for later stages. Duplicates will be removed at all the three stages of screening. Reasons for exclusion of studies will also be tabulated parallelly. The process from retrieval of studies to selection of articles for data synthesis will be illustrated in the form of a flowchart as per PRISMA-ScR guidelines. A workflow of the study processes from data retrieval to data synthesis has been provided in Figure 1.

**Figure 1:**
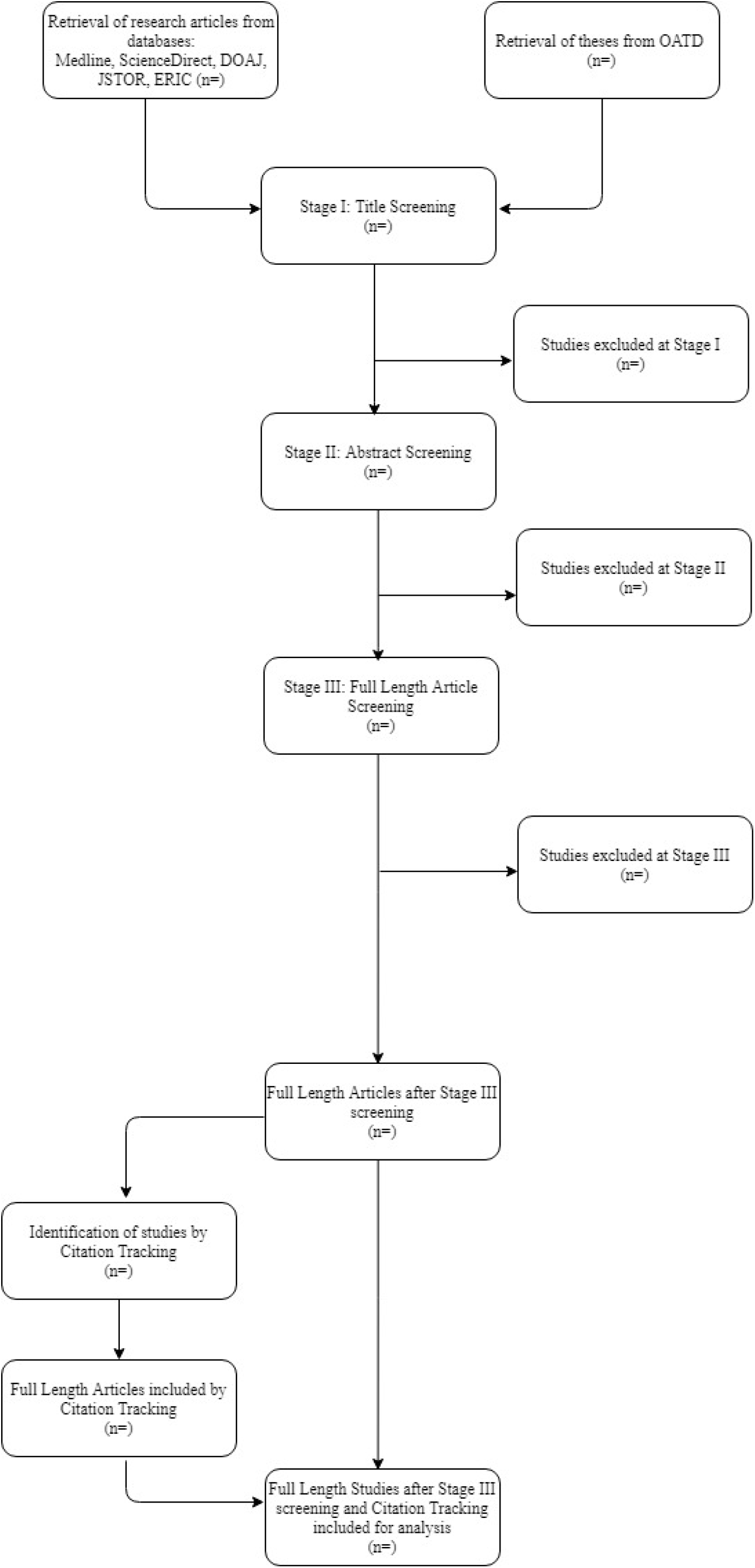
Flowchart of data flow:

### Data extraction

A data extraction template has been devised by the research team through iteration exercises for capturing information relevant to the research question(s). One reviewer will extract data from all the articles included in the study. A second reviewer will evaluate 10% of the articles randomly drawn from the master list, for quality control. Data extraction template has been exemplified in the Appendix II.

Extraction of data will be carried out in MS Excel 2019 or other similar electronic data spreadsheet. The data extracted by two different reviewers during quality control will be assessed for agreement using kappa statistics. The overall kappa values of greater than 0.8 will be considered as high level of agreement. For further quality assurance, we will employ lot quality assessment sampling, where the whole data extraction exercise would be repeated from scratch if more than 20% difference in agreement is observed in 10% of the total studies randomly selected for quality check.

### Data presentation

The key characteristics of studies analysed will be represented using simple descriptive statistics. Thematic analysis approach will be employed to analyse the extracted and tabulated data. Document analysis approach of content analysis will be used to synthesize evidence. The extracted data will be collated and discussed by all the reviewers for synthesis. Deductive approach will be used to identify the factors associated with salt consumption pertaining to the central themes of the study, namely:

a. Risk perception (severity and susceptibility): regarding sources of dietary salt; awareness regarding the associated illnesses and risk of developing these illnesses up on excessive salt consumption.
b. Practices: dietary practices at home, workplace and other places that contribute to excessive/uncontrolled salt intake; environmental factors responsible for these practices.
c. Barriers: perceptions, beliefs and/or physical barriers in reducing dietary salt intake; gaps which inhibit translation of knowledge into practice.

If deemed necessary during the analysis stage, inductive approach will be used to identify newly emerging themes. Three thematic maps will be designed to represent the results of thematic analysis, one based on Health Belief Model, another on Transtheoretical Model and a third composite map of both the models.

## Data Availability

We shall make all the data available after the scoping review is completed.

---

**Appendix I:**
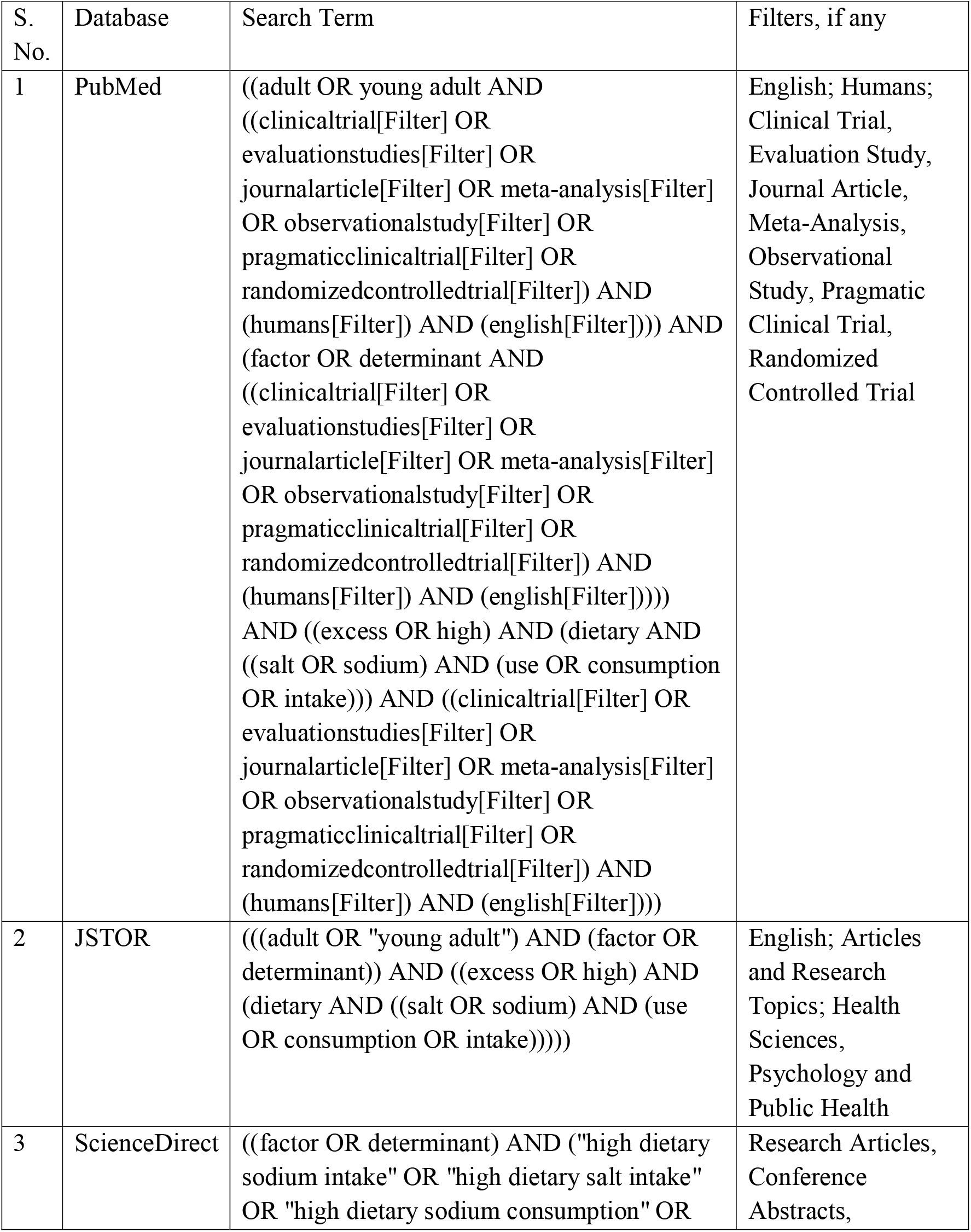
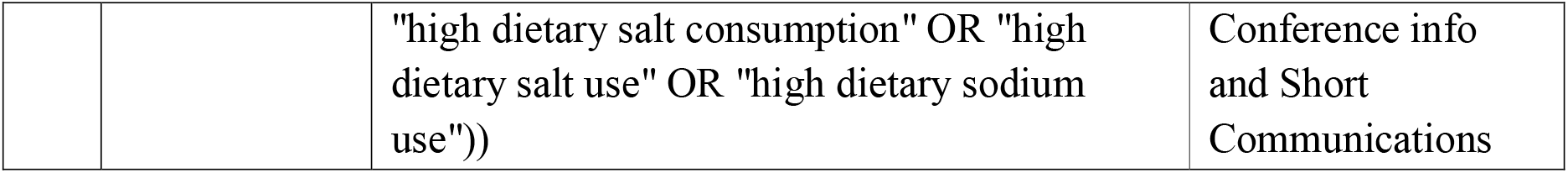
Final Search Strategy for Data Retrieval from MEDLINE:

**Appendix II:**
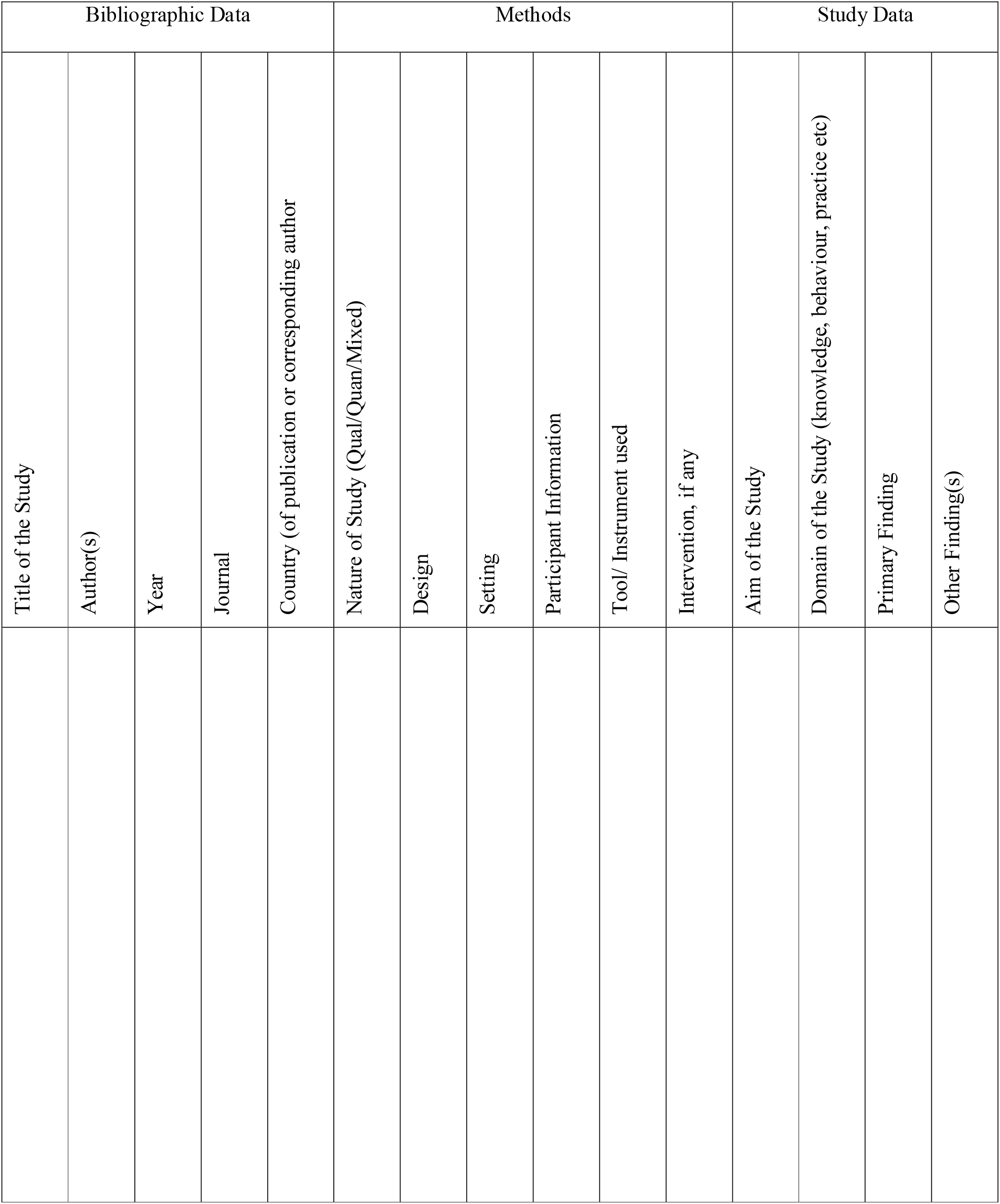
Data extraction template:

